# Evaluation of Molecular-based Methods for the Detection and Quantification of *Cryptosporidium* spp. in Wastewater

**DOI:** 10.1101/2024.03.01.24303624

**Authors:** Oumaima Hachimi, Rebecca Falender, Gabriel Davis, Rispa Vranka Wafula, Melissa Sutton, June Bancroft, Paul Cieslak, Noah Lininger, Christine Kelly, Devrim Kaya, Tyler S. Radniecki

## Abstract

*Cryptosporidium*, a eukaryotic protozoan parasite, poses a significant public health risk as a cause of waterborne disease worldwide. Clinical surveillance of cryptosporidiosis is estimated to be largely underreported due to the asymptomatic and mildly symptomatic infections, clinical misdiagnoses, and barriers to access testing. Unlike clinical surveillance, wastewater surveillance overcomes these limitations and could serve as an effective tool for identifying cryptosporidiosis at the population level. Despite its potential, the lack of standardized methods for *Cryptosporidium* spp. detection in wastewater challenges the comparability of studies. Additionally, the use of standard methods for the detection and quantification of *Cryptosporidium* spp. in surface waters may not be appropriate for wastewater samples due to the more complex composition of the wastewater matrix. Thus, in this study, we evaluated methods for concentrating *Cryptosporidium* oocysts from wastewater, extracting DNA, and detecting its genetic markers, using wastewater seeded with *C. parvum* oocysts. The evaluated concentration methods included electronegative membrane filtration, Envirocheck HV capsule filtration, centrifugation, and Nanotrap Microbiome particles. All methods except the Nanotrap Microbiome particles were conducted with or without additional purification via immunomagnetic separation. For DNA extractions, we tested the DNeasy Powersoil Pro kit and the QIAamp DNA Mini kit, while assessing the impact of bead beating and freeze-thaw cycles on DNA yield. Genetic detection was performed using qPCR targeting the *Cryptosporidium* 18S rRNA gene. Our results indicated that centrifugation yielded the highest oocyst recoveries (39-77%), followed by the Nanotrap Microbiome particles (24%), electronegative filtration with a phosphate buffered saline with 20% Tween 80 elution (22%), and Envirocheck HV capsule filtration (13%). IMS was found to be unsuitable due to interference from the wastewater matrix, significantly reducing recovery rates to 0.03 to 4%. DNA yields were highest with bead-beating pretreatment with either the DNeasy Powersoil Pro kit (314 gc/μL DNA) or the QIAamp DNA Mini kit (238 gc/μL DNA). In contrast, freeze-thaw pretreatment reduced DNA yields to under 92 gc/μL DNA, likely through DNA degradation. This study is amongst the first to compare different concentration methods, including Nanotrap Microbiome particles, and DNA extraction methods that can be utilized for *Cryptosporidium* spp. wastewater surveillance, highlighting the importance of method selection for accurate detection and quantification.

## 1. Introduction

*Cryptosporidium* spp. are eukaryotic protozoan parasites within the phylum Apicomplexa and subgroup coccidia (O’Donoghue, 1995). Unlike other parasites, *Cryptosporidium* spp. are intracellular and extracytoplasmic, undergoing a monoxenous life cycle where they complete their entire life cycle, both sexual and asexual stages, within a single host (Ghazy et al., 2015). They are distinguished by other several features including their small spherical size (diameter typically around 4 – 6 μm), minimal mitochondrial genome (∼9.2 Mbp), the ability to initiate self-infection in both zoonotic and anthroponotic potentials, their high resistance to disinfectants (up to 15,300 mg-min/L), and the ability to retain infectivity potential for up to several months outside of their hosts (CDC, 2021; Ghazy et al., 2015; Helmy & Hafez, 2022; Khan et al., 2018; King & Monis, 2007; Shields et al., 2008). To date, around 44 *Cryptosporidium* species have been identified, some of which are host-specific while others are ubiquitous in terms of host infectivity (Ryan et al., 2021).

*Cryptosporidium* stands as one of the most prominent waterborne enteric pathogens with substantial public health implications (Efstratiou et al., 2017; Gururajan et al., 2021)). After retroviruses, cryptosporidiosis infection is the second leading cause of both diarrhea and death (Khalil et al., 2018). Additionally, cryptosporidiosis ranks as the fourth leading cause of death from gastrointestinal diseases among young children under the age of five (Kotloff et al., 2013; Troeger et al., 2017). Consequently, *Cryptosporidium* has emerged as a significant etiology of long-term health issues, including malnutrition, stunted growth, and cognitive deficits in young children, as well as colon cancer in immunocompromised patients (Kotloff et al., 2019; Osman et al., 2017; Shirley et al., 2012).

Despite being a substantial public health concern and a nationally reportable disease, the CDC estimates that cryptosporidiosis routine clinical-based surveillance reports less than 2% of cases in the United States (CDC, 2021). Similar patterns are seen globally with cryptosporidiosis cases being underreported by an estimated two orders of magnitude or more (HALL et al., 2006; Zahedi et al., 2021). The underreporting of cryptosporidiosis cases and the underestimation of cryptosporidiosis prevalence have been attributed to asymptomatic and mildly symptomatic infections, barriers in access to testing, limited and voluntary testing by clinicians, as well as misdiagnosis due to symptoms resembling other gastrointestinal illnesses (O’Leary et al., 2021; Painter et al., 2015).

Additionally, stool microscopy, the golden standard method, is typically characterized by poor sensitivity and limited capacity to distinguish between *Cryptosporidium* species (O’Leary et al., 2021; van Lieshout & Roestenberg, 2015). In contrast, immunoassays exhibit high antigenic variation leading to inaccuracy in detecting all infecting *Cryptosporidium* species and genotypes (Danišová et al., 2018; Helmy et al., 2014; Uppal et al., 2014; Weitzel et al., 2006). Molecular techniques, on the other hand, offer a good alternative to both microscopy and antigen testing, however, they require specialized high-throughput laboratories and skilled technicians, resources that may not be available, particularly in cryptosporidiosis endemic countries (Meurs et al., 2017).

Given the documented underreporting of cryptosporidiosis, wastewater surveillance of *Cryptosporidium* spp. may provide an important supplemental surveillance technique to identify and monitor potential cryptosporidiosis outbreaks at the population level. Wastewater surveillance, also known as wastewater-based epidemiology, is the quantification of pathogens in wastewater to monitor disease burden in a community. It has been successfully applied as a reliable and non-invasive epidemiological tool for various pathogens including SARS-CoV-2, influenza, and RSV (Hughes et al., 2022; Layton et al., 2022; Wolfe et al., 2022). Cryptosporidiosis is another potential candidate for successful wastewater surveillance as symptomatic and asymptomatic individuals can excrete up to 10^10^ *Cryptosporidium* oocysts per bowl movement into wastewater, leading to wastewater concentrations as high as 60,000 oocysts per L being reported (Hamilton et al., 2018; Helmy & Hafez, 2022; Zahedi et al., 2021). Additionally, *Cryptosporidium* oocysts neither replicate nor decay rapidly in wastewater, leading to a reliable signal in the wastewater (Chalmers & Davies, 2010; B. J. King & Monis, 2007; Walker et al., 1998).

In general, the molecular detection and quantification of *Cryptosporidium spp.* in wastewater is a complex process that typically involves four steps: 1) representative sampling, 2) concentration of the oocysts, 3) extraction of DNA from the oocysts, and 4) quantification of the oocysts DNA by a PCR-based method. However, while *Cryptosporidium* has been detected in wastewater across the globe, there are currently no standardized concentration methods or molecular techniques to detect and quantify *Cryptosporidium* in wastewater (Zahedi et al., 2021).

For example, a variety of oocyst concentration methods have been used with filtration and centrifugation being the most common (Huang et al., 2017; L. Ma et al., 2019; Schmitz et al., 2018; Zahedi et al., 2018). Other less commonly used methods include polyethylene glycol precipitation (PEG) *(Pecson et al., 2022)*, calcium carbonate flocculation (Ma et al., 2016), aluminum sulfate flocculation (Gallas-Lindemann et al., 2013), skimmed milk flocculation *(Gonzales-Gustavson et al., 2017)*, glucose flotation (Lonigro et al., 2006), salt flotation (Wells et al., 2016), as well as formol-ether concentration (Lora-Suarez et al., 2016). Additionally, new wastewater concentration techniques, such as Nanotrap Microbiome particles, are entering the marketplace and offer the potential for rapid workflow and increased automation and throughput (Ceres Nanosciences, Inc. | United States, 2024). To date, there have been limited comparison studies as to how these different concentration methods, including the wastewater volumes used, affect oocyst recovery from wastewater (Montemayor et al., 2005).

Similarly, DNA extraction protocols can differ widely, especially in terms of different pretreatments to effectively disrupt the thick and robust *Cryptosporidium* oocyst wall along with commercial DNA extraction protocols. Pretreatments can either be mechanical such as bead-beating (*Elwin et al., 2012; Valeix et al., 2020*), freeze-thawing (Nichols et al., 2004; Lindergard et al., 2012), and sonication (Anceno et al., 2007; Adamska et al., 2011), or chemical using different lysis buffers (Paulos et al., 2016). Thus far, there have been limited comparison studies on how these different DNA extraction techniques affect *Cryptosporidium* gDNA recovery from wastewater samples (Mthethwa et al., 2022).

Molecular detection methods can also vary widely from qualitative methods of endpoint polymerase chain reaction (PCR), restriction fragment length polymorphism, and nested PCR, to quantitative methods of quantitative PCR (qPCR), droplet digital PCR (ddPCR), and digital PCR (dPCR) (Amar et al., 2004; Kitajima et al., 2014; Mthethwa et al., 2022; Pomari et al., 2019; Wiedenmann et al., 1998; Yang et al., 2014). Finally, the target gene for the molecular detection methods can vary from 18S small subunit ribosomal RNA (18S rRNA), actin gene, 60-kDa glycoprotein gene (GP60), 70-kDa heat shock protein (HSP70) to *Cryptosporidium* outer wall protein (COWP) gene (Gobet & Toze, 2001; Mthethwa et al., 2022; Shin et al., 2018; Yang et al., 2014; Zahedi et al., 2018). These differences in sample concentration, DNA extraction, and molecular detection methods for the detection of *Cryptosporidium* in wastewater result in highly variable detection limits, specificity, and recovery, making it difficult to compare between studies (King et al., 2015; Kitajima et al., 2014; Mthethwa et al., 2022; Ramo et al., 2017; Ward et al., 2002; Yamashiro et al., 2019).

Therefore, the aims of this study are: (i) characterize a *Cryptosporidium* qPCR assay for detection limits and specificity, (ii) evaluate eight DNA extraction protocols, consisting of two commercial kits (DNeasy Powersoil Pro kit and QIAamp DNA Mini kit) and two mechanical pretreatment steps (bead-beating and freeze-thawing), for effective DNA extraction of *Cryptosporidium* spp. from spiked influent wastewater samples, and (iii) evaluate the *Cryptosporidium* oocyst recovery rates for four different wastewater concentration methods, including centrifugation, electronegative membrane filtration, Envirocheck HV capsules filtration, immunomagnetic separation, and Nanotrap Microbiome particles, across various wastewater volumes (10, 30, 100, 500, and 1000 mL).

## 2. Materials and Methods

### 2.1 Wastewater Preparation

A bulk wastewater sample (∼30 L) was collected from the influent of the Corvallis, Oregon, USA wastewater treatment plant prior to primary treatment. The wastewater was transported on ice to the laboratory and stored at 4 °C until further use. The *C. parvum* oocysts used to spike wastewater samples were purchased from the *Cryptosporidium* Production Laboratory at the School of Animal and Comparative Biomedical Sciences, the University of Arizona.

### 2.2 Cryptosporidium qPCR Assay

The *Cryptosporidium* 18S rRNA qPCR assays (***Table 1***) were performed in triplicate on a Bio-Rad CFX Connect Real-Time PCR Detection System (Bio-Rad Laboratories, Hercules, CA). The reaction mixtures contained 10 μL of 2X PrimeTime Gene Expression Master Mix (Cat No. 1055770 IDT, Coralville, IA), 2 μL of the primers and probe solution, 5 μL of template DNA, and 3 μL of nuclease-free water to bring the final reaction volume up to 20 μL. The thermocycling conditions for the qPCR assays used are reported in ***Table.1***. Both threshold cycle values and starting quantities were automatically calculated by the Bio-Rad CFX Manager software.

**Table 1.**
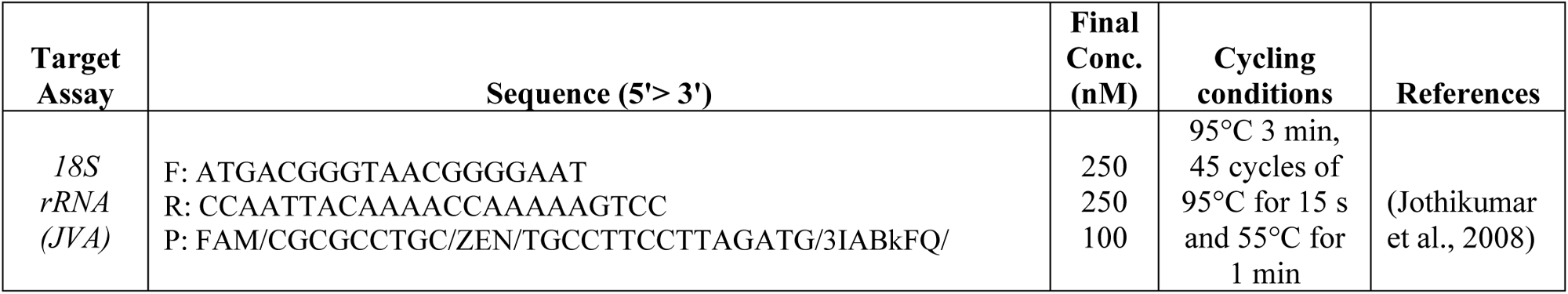
*Cryptosporidium* spp. primers and probe.

gBlock gene fragments of the target 18S rRNA gene (***Table 2***) was purchased from Integrated DNA Technologies (IDT, Coralville, IA) and was used to prepare 10-fold serial dilution qPCR standards curves ranging from 10^0^ to 10^6^ gene copies per reaction. Each qPCR run also included a no-template negative control (consisting of nuclease-free water) and a non-specific amplification negative control (consisting of *Escherichia coli* DNA).

**Table 2.**
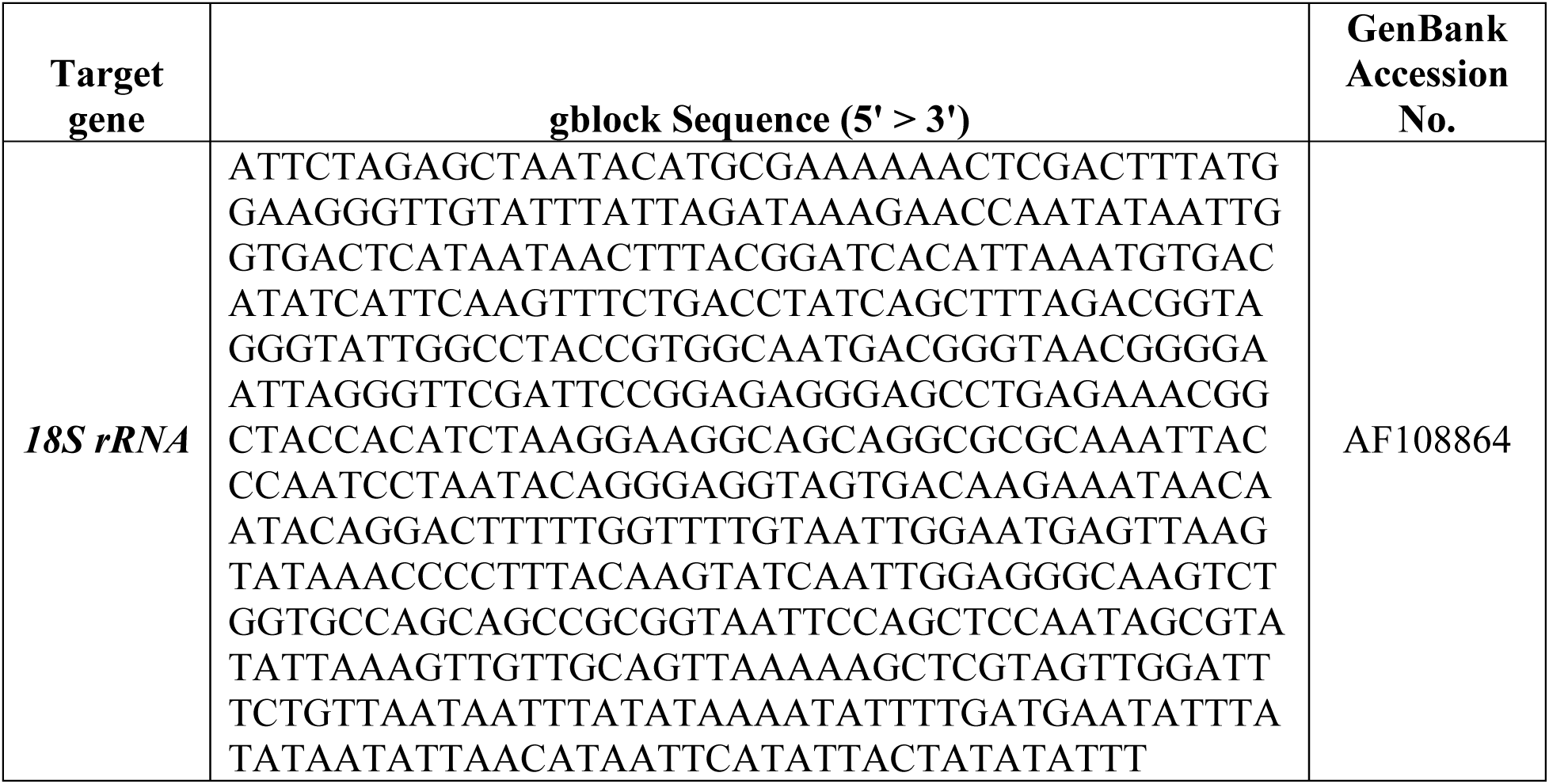
*Cryptosporidium parvum* gblock 18S rRNA gene fragment.

The sensitivity of the 18S rRNA qPCR assay was determined using its standard curve. The limit of blank (LOB) and limit of detection (LOD) of each assay were calculated from the assay standard curves using Equations 1 and 2, respectively, where σ is the calculated standard deviation (Forootan et al., 2017). The specificity of the primers and probe for the 18S rRNA qPCR assay was assessed through a BLAST analysis against the National Center for Biotechnology Information (NCBI) database.

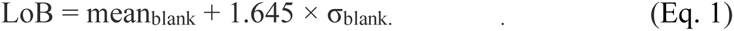

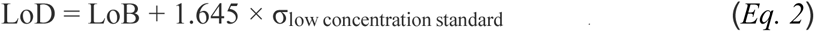

### 2.3 DNA Extraction

A total of eight DNA extraction protocols were conducted in triplicate on both wastewater that was seeded with 1 x 10^5^ *C. parvum* oocysts and wastewater that was not seeded (to account for background levels of *Cryptosporidium* naturally in the wastewater). Thirty milliliters of the wastewater samples were filtered through an electronegative mixed cellulose ester membrane filter (catalog no. 7141-104, Whatman, Buckinghamshire, U.K), as described in *Section 2.4.2*, and were stored at 4 °C for less than 24 h until DNA extraction could occur.

Each DNA extraction protocol utilized one of two commercial DNA extraction kits, the QIAamp DNA Mini kit (Qiagen, Germany) or the DNeasy Powersoil Pro kit (Qiagen, Netherlands), per the manufacturer’s instructions (***Table 3***). The DNA extraction kits were used either independently (*M1 & M2*) or in combination with bead-beating (*M3 & M4*), freeze-thawing (*M5 & M6*), or both (*M7 & M8*). Samples subjected to bead-beating were homogenized by a BioSpec Mini-Beadbeater 16 (BioSpec Products, Inc., Bartlesville, OK) for 2 min. After cooling on ice for 2 min, the samples were centrifuged at 12,000 x g for 1 min and DNA was extracted from 250 μL of the supernatant. Samples subjected to freeze-thaw cycles were frozen in liquid nitrogen for 10 min and thawed in an 80 °C water-bath for 5 min for five freeze-thaw cycles before DNA was extracted from 250 μL aliquot of the lysate. Finally, samples subjected to both pretreatment methods underwent bead-beating first, followed by freeze-thawing.

**Table 3.**
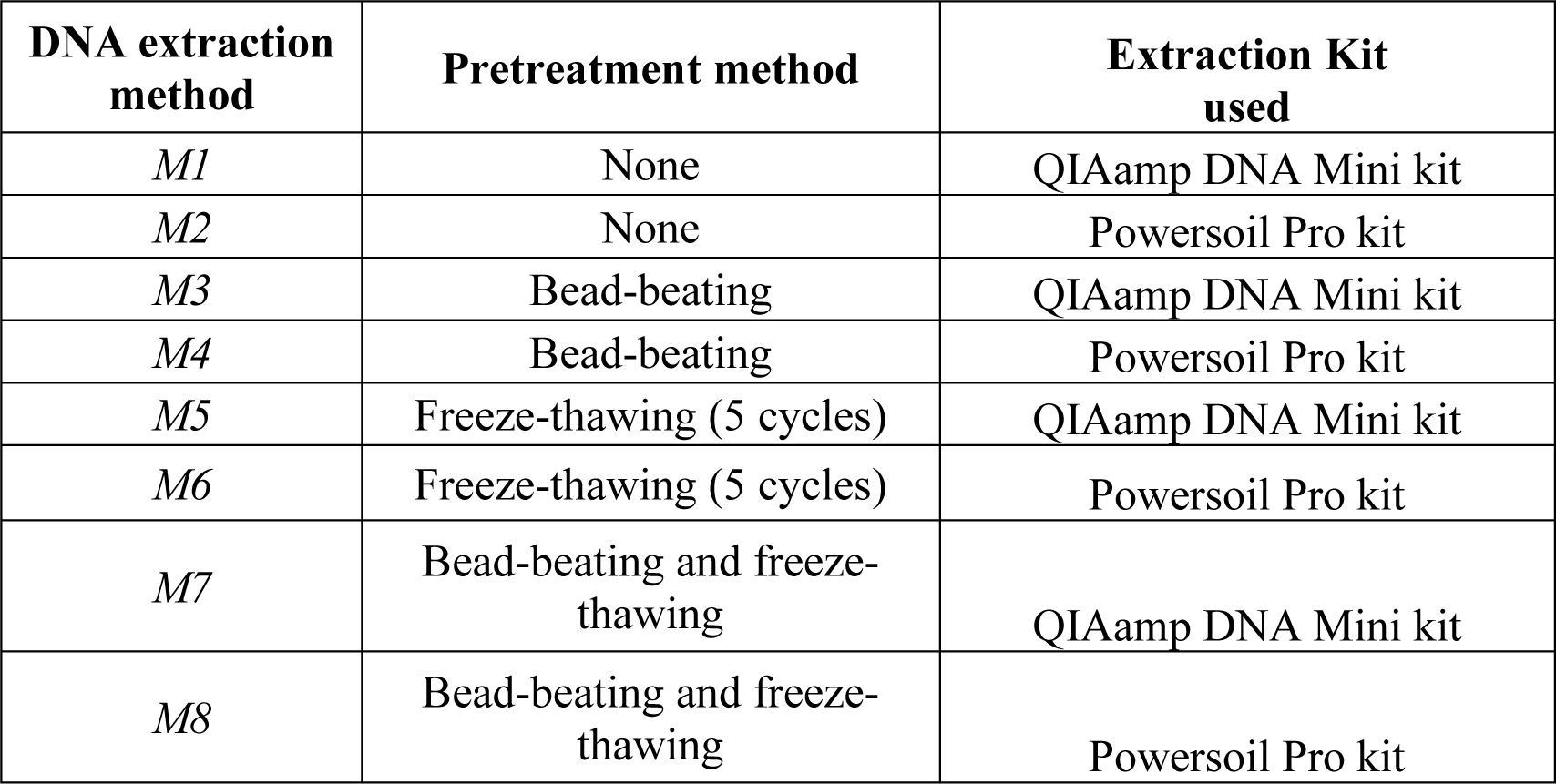
DNA extraction methods.

| DNA extraction method | Pretreatment method | Extraction Kit used |
| --- | --- | --- |
| *M1* | None | QIAamp DNA Mini kit |
| *M2* | None | Powersoil Pro kit |
| *M3* | Bead-beating | QIAamp DNA Mini kit |
| *M4* | Bead-beating | Powersoil Pro kit |
| *M5* | Freeze-thawing (5 cycles) | QIAamp DNA Mini kit |
| *M6* | Freeze-thawing (5 cycles) | Powersoil Pro kit |
| *M7* | Bead-beating and freeze-thawing | QIAamp DNA Mini kit |
| *M8* | Bead-beating and freeze-thawing | Powersoil Pro kit |

After DNA extraction, the concentration of the *Cryptosporidium* 18S rRNA gene was quantified via qPCR, as described in *Section 2.2*, and converted into gene copies/μL DNA (***Equation 3***). To calculate the recovered seeded *Cryptosporidium* 18S rRNA gene concentration, the concentration of *Cryptosporidium* 18S rRNA genes from the unseeded wastewater samples was subtracted from the concentration of the *Cryptosporidium* 18S rRNA genes from the seeded wastewater samples.

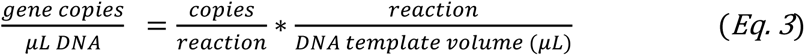

### 2.4 Wastewater Concentration

Wastewater aliquots of 10-, 30-, 100-, 500-, and 1000-mL volumes were seeded with 1 × 10^5^ *C. parvum* oocysts and thoroughly mixed before undergoing a concentration method. Unseeded wastewater aliquots were also concentrated. After the concentration method had been completed, all samples were stored at 4 ^°^C for less than 24 h until DNA extraction using Method 4 (*M4*), as described in *Section 2.3*, could occur.

*Cryptosporidium* concentrations were determined via qPCR targeting the 18S rRNA gene, as described in *Section 2.2*. The qPCR results were converted into *Cryptosporidium* oocysts concentrations (***Equation 4***). The percent recovery of each concentration method was determined by dividing the recovered seeded *Cryptosporidium* oocyst mass (***Equation 5***) by the mass of *Cryptosporidium* oocysts seeded into the wastewater (***Equation 6***).

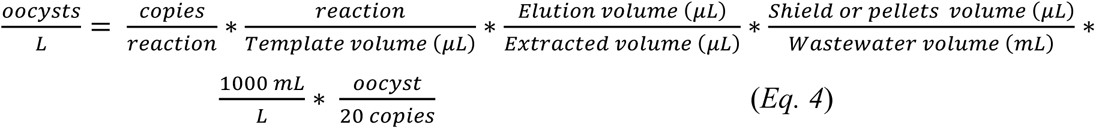

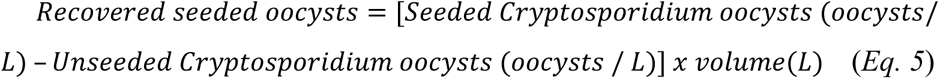

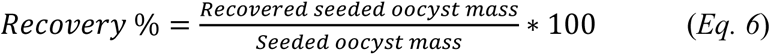

#### 2.4.1 Centrifugation

Wastewater aliquots were centrifuged at *2000 x g* for 30 min in a swinging-bucket rotor using either 50 mL or 250 mL sterile conical tubes, for wastewater volumes of 30 mL or greater than 30 mL, respectively. The supernatant was carefully removed leaving 5 mL of supernatant for every 0.5 mL of pellet (USEPA Method 1693, 2014). For wastewater volumes greater than 250 mL, multiple 250 mL sterile conical tubes were used, and the pellets were combined into a single 5 mL suspension before DNA extraction occurred.

#### 2.4.2 Electronegative Filtration (HA)

Wastewater aliquots of 30 mL and 100 mL volumes were filtered through a 0.45 μm electronegative mixed cellulose ester membrane filter (catalog no. 7141-104, Whatman, Buckinghamshire, U.K). After filtration, the filters were placed in a 2 mL tube containing 0.7 mm garnet beads and 1 mL of DNA/RNA Shield (Zymo Research, Irvine, CA) and stored at 4 °C for less than 24 h until DNA extraction could occur. To reduce clogging, the filtration of the 100 mL volumes was split into four filtration steps, with each filter processing 25 mL of wastewater. The four filters were placed sequentially into the same 2 mL beaded tube with each filter undergoing bead beating before being removed and the next filter was added.

In addition, PBST elution buffer (Phosphate Buffered Saline with 20% Tween 80) was evaluated as a potential washing step to enhance *Cryptosporidium* oocyst recoveries. In this case, 30 mL wastewater samples were filtered using the above protocol and placed in a 2 mL tube containing 0.7 mm garnet beads and 1 mL of DNA/RNA Shield. However, prior to bead beating, the filter was removed from the DNA/RNA Shield containing tube and placed in a new 2 mL tube containing 0.7 mm garnet beads and 1 mL of PBST elution buffer. Bead-beating commenced using the PBST elution buffer containing a beaded tube.

#### 2.4.3 Envirocheck Filtration

A modified version of USEPA Method 1693 concentration by filtration was conducted by filtering 500, or 1000 mL of wastewater through an Envirocheck polyester HV capsules (catalog no. 12098, Pall Corporation, New York, USA). Prior to elution, the capsules were rinsed with 5% sodium hexametaphosphate (NaHMP). Subsequently, the materials retained on the filters were eluted using PBST elution buffer. The resulting eluate was concentrated via centrifugation.

### 2.5 Immunomagnetic Separation (IMS)

To potentially increase the specificity of oocyst concentration and purification protocols, an immunomagnetic separation (IMS) procedure was performed on wastewater samples previously concentrated by either centrifugation (Section 2.4.1), electronegative filtration (Section 2.4.2) or Envirocheck filtration (Section 2.4.3). Dynabeads anti-*Cryptosporidium* beads were used to magnetically retain *Cryptosporidium* oocysts from each sample while discarding extraneous materials following manufacturer’s instructions (catalog no. 73011, Applied Biosystems, Waltham, MA, USA). The recovery efficiencies of the IMS method alone were evaluated by seeding *C. parvum* oocysts into 10 mL of either nuclease-free water or wastewater and directly concentrating and eluting, via two acid dissociations, the samples per manufacturer’s instructions.

### 2.6 Nanotrap Microbiome B Particles

Nanotrap Microbiome B particles (Ceres Nanosciences, Manassas, VA, USA) were used to capture and concentrate *Cryptosporidium* oocysts from 10 mL wastewater samples, per manufacturer’s instructions. In brief, Nanotrap Microbiome B magnetic particles and enhancement reagent 3 (ER3) were added to the wastewater samples to initiate affinity-based binding. Once the affinity-based binding was complete, the particles were magnetically separated from extraneous supernatant and eluted in a lysis buffer. DNA was directly extracted using the recommended accompanying MACHEREY-NAGEL DNA kit (Cat no. 744220.1, MACHERY-NAGEL, Düren, Germany), per manufacturer’s instructions.

### 2.7 Statistical analysis

Statistical analysis was conducted in Microsoft Excel. To determine statistical significance, student t-tests were applied to the data sets, two-tailed p values were calculated, and p < 0.05 was accepted as statistically significant.

## 3. Results and Discussion

### 3.1 Comparison of Cryptosporidium qPCR Assays

Selecting a qPCR assay capable of detecting all *Cryptosporidium* species and genotypes presents a significant challenge in accurate identification and quantification of this pathogen in wastewater samples. An effective approach involves targeting highly conserved areas of the *Cryptosporidium* genome (*e.g.,* the 18S rRNA gene) which contains a high copy number of interspecific polymorphisms in all species and genotypes (Roellig & Xiao, 2020). The *Cryptosporidium* 18S rRNA gene stands out as the most commonly used genetic target due to its low levels of intraspecific variation as well as its increased theoretical sensitivity as a multi-copy gene (O’Leary et al., 2021; Xiao, 2010). Therefore, our study targeted the *Cryptosporidium* 18S rRNA gene using primers and probe that were originally designed for *Cryptosporidium* spp. clinical surveillance from stool samples (Table 1).

To assess the assay’s sensitivity, the limit of blank (LOB) and limit of detection (LOD) were determined. In total, 99 qPCR reactions were run, encompassing no-template controls and 10-fold serially diluted standards ranging between 10^0^ to 10^6^ gene copies per reaction. The calculated LOB and LOD of the 18S rRNA assay were 0.0 and 2.91 gene copies per reaction, respectively. A specificity analysis using NCBI BLAST confirmed that the 18S rRNA assay could detect all *Cryptosporidium* spp., including C. *hominis*, C. *parvum*, C. *canis*, C. *andersoni*, C. *bovis*, C. *ubiquitum*, C. *meleagridis*, C. *muris*, C. *felis*, C. *baileyi*, C. *cuniculus*, C. *wrairi*, C. *erinacei*, and many other genotypes including horse, chipmunk, skunk, mink, and ferret genotypes.

### 3.2 Comparison of DNA Extraction Protocols

The detection and quantification of *Cryptosporidium* spp. in environmental samples highly relies on DNA extraction among other factors (Hawash, 2014). The main challenge in extracting DNA from *Cryptosporidium* spp. and other parasitic protozoa resides in breaking their robust thick outer wall (Samuelson et al., 2013). Prior research has indicated the necessity of pretreatments, either mechanical or chemical, to break this barrier and enhance DNA recoveries (Valeix et al., 2020).

To determine the most effective DNA extraction protocol for enhancing *Cryptosporidium* DNA recovery in wastewater, our study evaluated the DNA yields of eight different protocols which used combinations of two pretreatments (freeze-thaw and bead beating) and two commercial extraction kits (QIAamp DNA mini kit and DNeasy Powersoil Pro kit). Both pretreatments and commercial extraction kits have been previously utilizing in *Cryptosporidium* quantification from wastewater (Elwin et al., 2012; Lindergard et al., 2003; Mthethwa et al., 2022; Valeix et al., 2020). However, the specific effectiveness of these approaches in extracting *Cryptosporidium* DNA, both individually and in combination, had not been thoroughly explored.

Regardless of the extraction method employed, the 18S rRNA gene was quantifiable in all samples, demonstrating 100% qPCR positivity (***Figure 1***). Furthermore, in the absence of pretreatment, samples extracted using both the QIAamp DNA Mini kit and the Powersoil Pro kit resulted in comparably similar DNA yields (p > 0.05) with an average of 3.43 × 10^1^ (± 1.95) gene copies/μL and 3.26 × 10^1^ (± 2.03) gene copies/μL, respectively (M1 & M2). The bead-beating pretreatment (M3 & M4) drastically improved DNA yields by an order of magnitude, however, there were notable differences between the two extraction kits (p < 0.05). The bead-beating pretreatment increased the DNA yield to 3.14 × 10^2^ (± 3.61) gene copies/μL for the Powersoil Pro kit and to 2.38 × 10^2^ (± 2.52) gene copies/μL for the QIAamp DNA Mini kit. In contrast, the freeze-thawing pretreatment only increased the DNA yield of the QIAamp DNA Mini kits (M5) by roughly a factor of 2, up to 6.08 × 10^1^ (± 1.10) gene copies/μL and decreased the DNA yield of the Powersoil Pro kit (M6) by roughly a factor of 2, down to 1.74 × 10^1^ (± 2.84) gene copies/μL. Incorporating bead-beating prior to freeze-thawing significantly increased DNA yields (p < 0.05) for both kits (M7 and M8). However, once again, the QIAamp DNA Mini kits had a higher DNA yield, 9.19 × 10^1^ (± 2.82) gene copies/μL, compared to the Powersoil Pro kits, 4.46 × 10^1^ (± 1.06) gene copies/μL.

**Figure 1:**
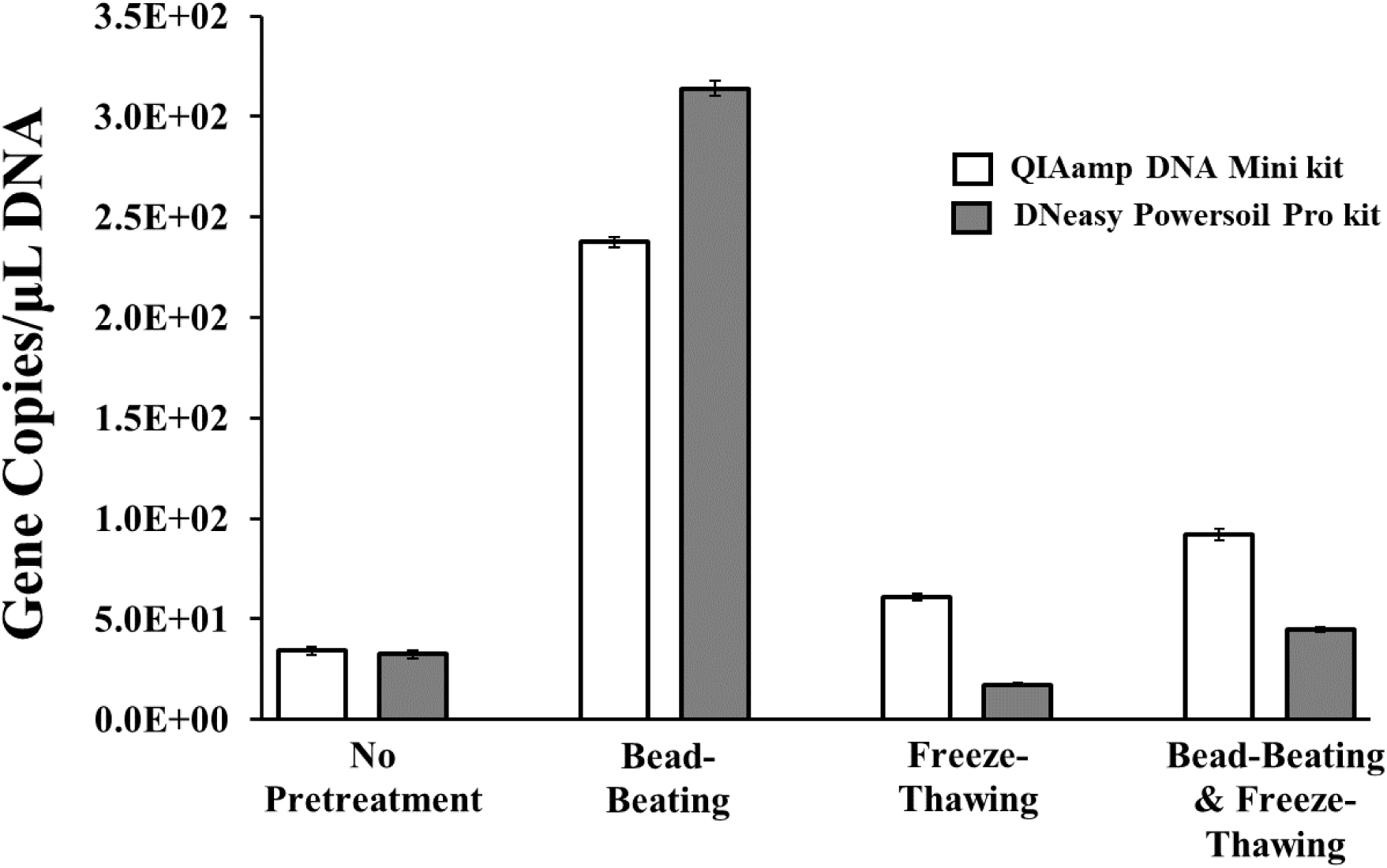
*Cryptosporidium* DNA yields (gene copies/μL of DNA) from either the QIAamp DNA mini kit (□) or the DNeasy Powersoil Pro kit (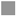) after a variety of pretreatment steps. Error bars represent standard deviation intervals.

Thus, bead-beating, emerged as the most effective method for rupturing the tough structure of the oocyst wall while also preserving genomic material by not causing excessive DNA shearing or hydrolysis. This finding aligns with other studies investigating *Cryptosporidium* and other parasites across various matrices (*Babaei et al., 2011; Amoah et al., 2019; Temesgen et al., 2020; Shipley et al., 2022*). Compared to other pretreatments, bead-beating not only improved DNA extraction but also enhanced PCR sensitivity, offering a rapid, cost effective, and straightforward approach for sample preparation (Elwin et al., 2014; Lindergard et al., 2003; Scharf et al., 2020; Shipley et al., 2022).

The lower DNA yields from freeze-thaw pretreatments could be due to DNA degradation, as has been reported in other studies using freeze-thaw pretreatment for DNA extraction from *Cryptosporidium* oocysts and other protozoa and parasites (Babaei et al., 2011; Mthethwa et al., 2022). Additionally, a reduction in detection sensitivity has been noted as a drawback associated with freeze-thaw cycles. *Lindergard et al. (2003)* reported that the highest sensitivity of detecting *C. parvum* and *T. gondii* oocysts was achieved using a single freeze-thaw cycle and that the DNA detection sensitivity and concentration decreased with each additional cycle due to DNA degradation (Lindergard et al., 2003). Furthermore, freeze-thaw pretreatments have been associated with the increased release of PCR inhibitors (D’Alessandro et al., 2007).

Incorporating a bead-beating step prior to the freeze-thaw cycles did result in improved DNA yields (p < 0.05). This is in alignment with other studies in which these two pretreatments demonstrated more satisfactory results for DNA extraction from *Cryptosporidium* oocysts and other protozoa compared to solely using freeze-thaw pretreatment (Babaei et al., 2011; Hamedi et al., 2016; Mthethwa et al., 2022). However, the DNA yields from combined pretreatments were still lower than those from bead-beating alone, indicating significant DNA loss due to degradation during the freeze-thaw process.

In contrast, the Powersoil Pro kits produced lower DNA yields than the QIAamp DNA Mini kit when a freeze-thaw pretreatment was applied, regardless of whether bead-beating was performed first. While some studies have shown that freeze-thaw pretreatment increased DNA yields when used with the QIAamp kit (Adamska et al., 2011; Xin et al., 2021), others have indicated that freeze-thaw cycles led to DNA degradation when used with the DNeasy Powersoil Pro kit (Portnoy & Baxter, 2023). This disparity could stem from the unique characteristics of each extraction kit. Both kits use lysis buffers, but the QIAamp kit also includes enzymatic lysis (Proteinase K) while the Powersoil Pro kit utilizes an additional physical disruption step through a 10-minute homogenization step. This additional homogenization step following freeze-thaw pretreatment exacerbated DNA degradation, likely through physical disruption of the DNA.

The results of this study also demonstrated that the choice of pretreatment will affect the efficiencies of the DNA kits. For instance, while there is no significant difference (p > 0.05) in DNA yields between the DNA kits when no pretreatment is applied, the Powersoil Pro Kit produced higher DNA yields than the QIAamp DNA Mini kit when bead-beating pretreatment was conducted. This can be explained by the higher sensitivity and inhibitor removal technology attributed to this kit, as well as its associated 10-minute homogenization step (Amoah et al., 2019; Feng et al., 2023; Shipley et al., 2022).

### 3.3 Comparison of Wastewater Concentration Protocols

Wastewater concentration is a critical step in the molecular detection of *Cryptosporidium* spp. and other protozoa within environmental matrices. The choice of concentration method can have significant impacts on the sensitivity and efficiency of downstream methods (*e.g.*, DNA extraction, PCR, and genomic sequencing), and can influence the observed microbial diversity within wastewater samples (Zahedi et al., 2021). Reported concentration methods for *Cryptosporidium* analyses include centrifugation, membrane filtration, and hollow fiber filtration (*e.g.*, Envirocheck). IMS is also often used to increase the specificity of the concentration protocol, remove additional impurities, and increase the concentration of oocysts in the sample (Lowery et al., 2000). Finally, Ceres Nanoscience’s Nanotrap® Microbiome Particles have been used to concentrate viral and bacterial pathogens in wastewater and may also be effective for *Cryptosporidium*, although it remains largely untested (*Ceres Nanosciences, Inc. | United States*, 2024). While there are numerous concentration methods available for *Cryptosporidium* spp., they have generally been developed and validated for concentrating oocysts from surface waters and not wastewater (Hassan et al., 2021). Given the lack of standardization and comparison of these methods for wastewater samples, our study evaluated these concentration protocols at different wastewater volumes, spiked with *Cryptosporidium* oocysts, to determine their yields of recovery for *Cryptosporidium* oocysts.

All the concentration methods yielded quantifiable recoveries of *Cryptosporidium* oocysts. However, the percent recoveries varied considerably based on the concentration method and wastewater volume (***Figure 2***). For instance, the filtration-based protocols (*i.e.*, electronegative membrane and Envirocheck) had statistically lower percent recoveries (p < 0.05) than the centrifugation and Nanotrap Microbiome particle protocols at comparable wastewater volumes. The lower performance of the filtration-based concentration protocols may be attributed to the inefficient dislodging of *Cryptosporidium* oocysts from the filters.

**Figure 2:**
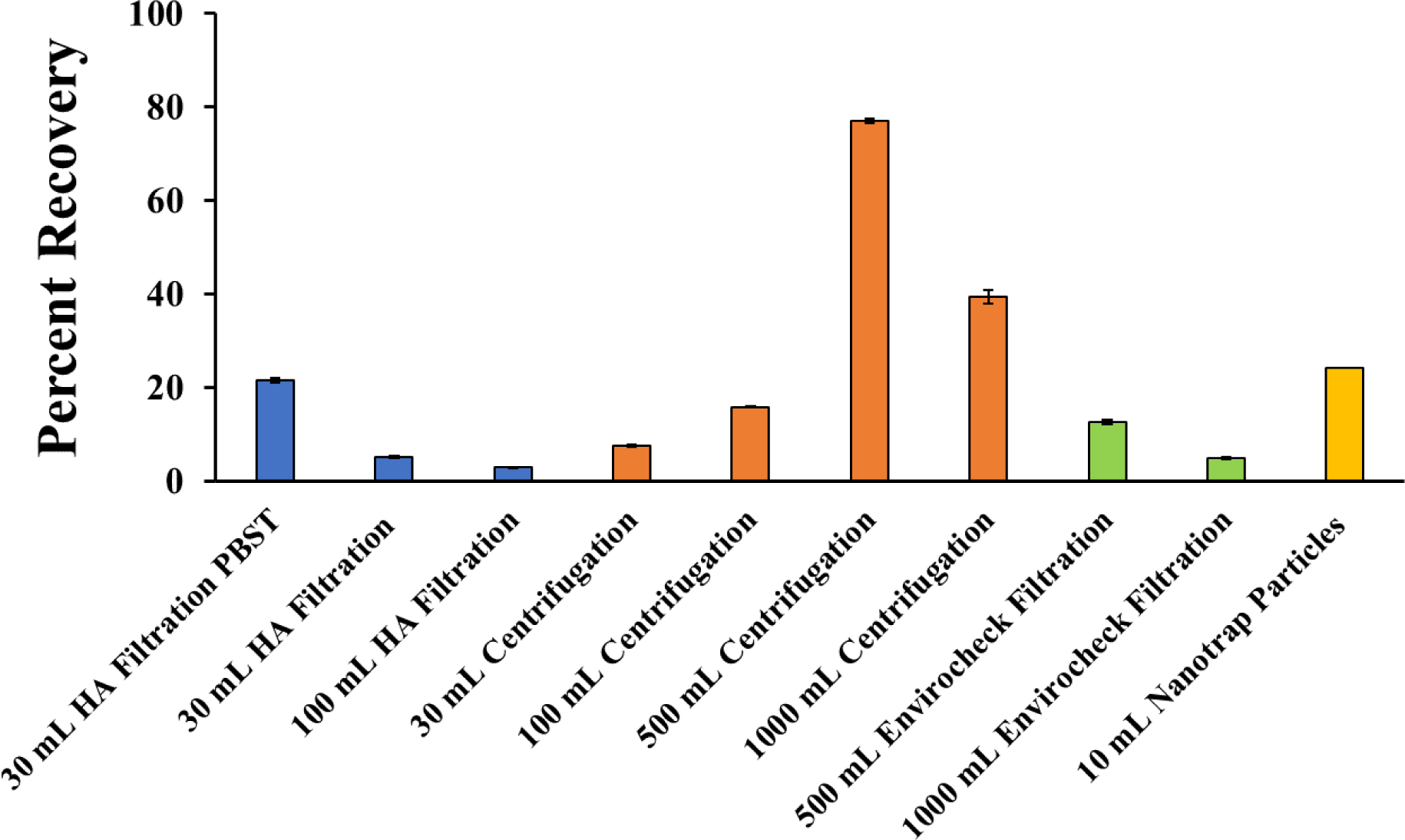
Percent recoveries of *Cryptosporidium* oocysts by various concentration methods at various wastewater volumes. Error bars represent standard deviation intervals.

For instance, the use of an electronegative membrane filter with a pore size of 0.45 μm should guarantee that all of the larger *Cryptosporidium* oocysts, with an average diameter of 4 - 6 μm, are removed by size exclusion (Fayer & Ungar, 1986). Yet, this method resulted in some of the lowest *Cryptosporidium* oocyst recoveries. However, when these filters were subjected to a PBST wash, the *Cryptosporidium* oocyst percent recoveries significantly increased (p < 0.05) from 5.1 ± 0.32% to 21.5 ± 0.39 %.

PBST contains Tween 80, a surfactant that can help dislodge solids from surfaces, and PBST washing is a standard step in the Envirocheck filtration protocol to help increase the dislodging of solids from the hollow fiber filters (USEPA Method 1693, 2014). As such, when PBST was used during the bead-beating step of the electronegative membrane filtration protocol, it helped to dislodge *Cryptosporidium* oocysts that were stuck to the electronegative filter. Based on the observed improvement in *Cryptosporidium* oocyst recoveries, it is recommended that a surfactant elution buffer, such as PBST, be included when conducting bead beating of electronegative membrane filters.

Unlike electronegative membrane filtration, the Envirocheck filtration protocol does not rely on strict pore-size exclusion but rather depth filtration and capture (Matheson et al., 1998), which allows the processing of larger volumes of water. When filtering 500 mL of wastewater, the *Cryptosporidium* oocyst percent recovery was higher (p < 0.05), 12.5 ± 0.45%, than that observed for electronegative membrane filtration at 30 mL without PBST, likely due to the inclusion of PBST in the Envirocheck filtration protocol. However, this percent recovery was lower than that observed for electronegative membrane filtration at 30 mL with PBST (p < 0.05). Additionally, the percent recovery decreased from 12.5 ± 0.45 % to 4.9 ± 0.22% when the wastewater volume increased from 500 mL to 1,000 mL. These findings suggest that the Envirocheck filter was also being clogged by wastewater solids and this clogging was preventing the elution of oocysts from the filter. It is important to note that this method (USEPA 1693, 2014) was originally developed for surface water filtration and the standard method does not recommend the use of hollow fiber filters with high turbidity samples, but rather centrifugation (USEPA Method 1693, 2014).

Overall, centrifugation-based concentration protocols yielded higher oocysts recoveries starting at 7.5 ± 0.28 % and 15.8 ± 0.11% when 30 mL or 100 mL of wastewater was processed, respectively, up to 77.01 ± 0.39% and 39.3 ± 1.4% when 500 mL or 1,000 mL of wastewater was processed, respectively. The increased percent recoveries with increasing wastewater volumes may be due to the increased mass of solids in the centrifuge tubes. As the wastewater solids pellet, they are likely to capture *Cryptosporidium* oocyst and help collect them in the pellet. More solid mass in the centrifuge tube results in a higher likelihood that the *Cryptosporidium* oocyst will be collected at the bottom of the centrifuge tube. A similar study used centrifugation as a concentration method of *Cryptosporidium* spp. in wastewater and found an oocyst recovery of 53.63%, when centrifuging 1L of influent wastewater at 3500 rpm (max 4 x 340 g) for 10 min (Mthethwa et al., 2022).

The Nanotrap Microbiome particles are porous magnetically functionalized hydrogels containing binding receptors with an affinity for biological materials (*Ceres Nanosciences, Inc. | United States*, 2024). The Nanotrap Microbiome particle protocol has been successfully used to quantify viral and bacterial pathogens in wastewater (Ahmed et al., 2023; Boehm et al., 2023). In this study, the Nanotrap Microbiome particle protocol produced similar results to the electronegative membrane filtration with PBST protocol at 24.1% and 21.5 ± 0.57%, respectively (p > 0.05). This agrees with previous work that found similar performances between the Nanotrap Microbiome particle protocol and electronegative membrane filtration for SARS-CoV-2 (Liu et al., 2023).

A key advantage of the Nanotrap Microbiome particle protocol is its ability to work in an automated workflow, due to the use of magnetic beads and a small sample volume of 10 mL. However, the percent recoveries from the Nanotrap Microbiome particle protocol were lower than the percent recoveries from the centrifugation protocol at the 500- and 1,000-mL volumes (p < 0.05). The reason for the lower percent recoveries is uncertain but may stem from complex wastewater matrix interference with the particles’ binding affinity for *Cryptosporidium*.

Finally, in an effort to enhance the specificity of *Cryptosporidium oocyst* detection in wastewater samples, immunomagnetic separation (IMS) was performed on wastewater samples previously concentrated by centrifugation or filtration. Increased specificity of the *Cryptosporidium* oocyst concentration protocol would potentially increase the sensitivity and data quality of downstream processes, including qPCR and next-generation sequencing. Although IMS has shown higher specificity and oocysts recovery in other studies on surface water and food matrices (*McCuin et al., 2001; McCuin et al., 2005; Plutzer et al., 2010; Silva et al., 2020*), our findings indicate that this method yielded lower recoveries in wastewater samples, with the majority of recoveries being less than 0.4% (***Figure 3***).

**Figure 3:**
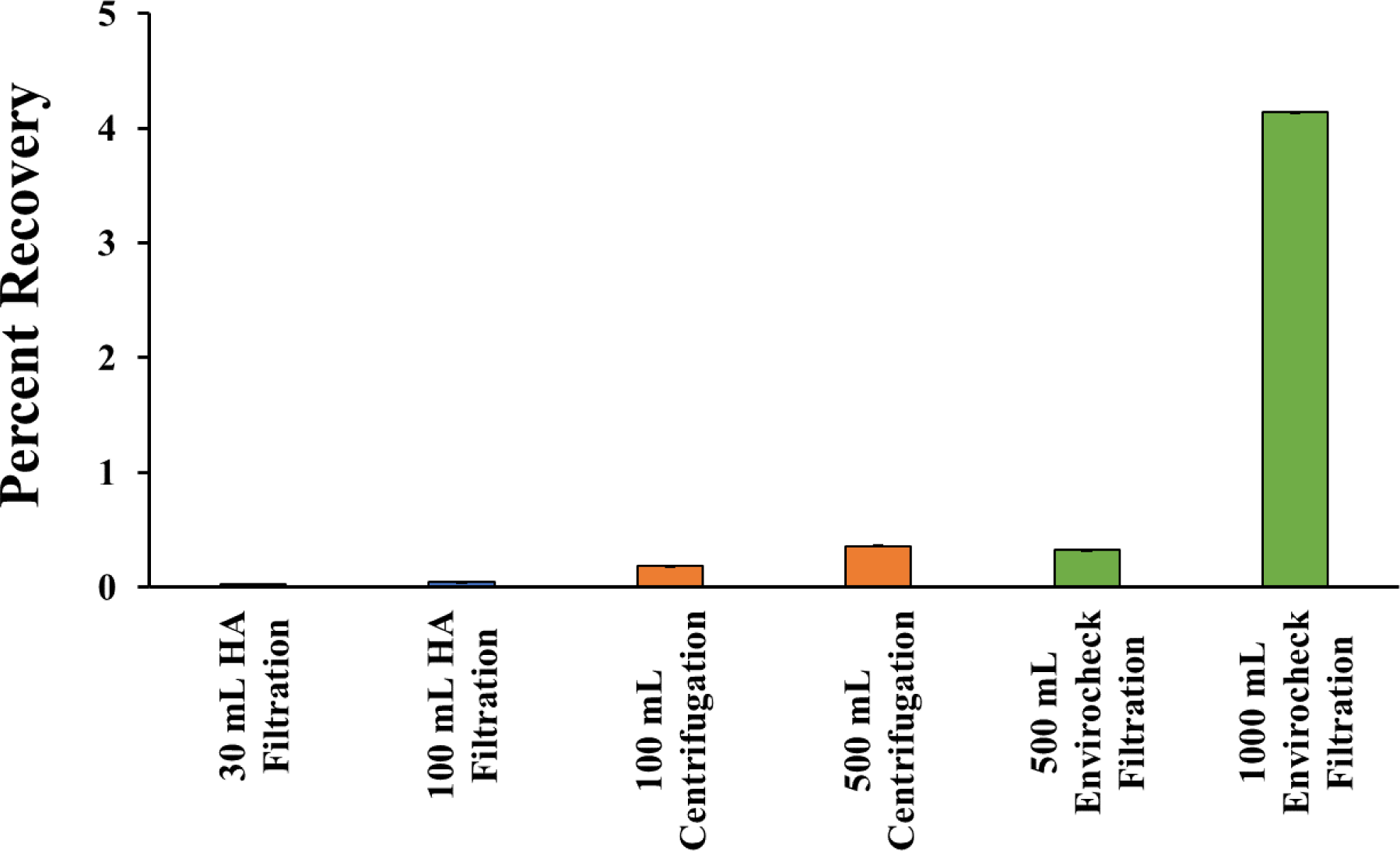
*Cryptosporidium* oocyst percent recoveries post IMS concentration protocol. Error bars represent standard deviation intervals.

When used under pristine conditions (*i.e.*, nuclease-free water spiked with *Cryptosporidium* oocysts), the percent recovery from the IMS protocol was 100% (data not shown). Thus, it was the wastewater matrix diversity and complexity that might have inhibited the performance of this method (McCuin & Clancy, 2006). Higher turbidity, particulate matter, suspended solids, organic and inorganic substances as well as fats and oils, and the other inhibiting substances present in wastewater could then interfere with the antibody binding to the *Cryptosporidium* oocysts leading to non-specific binding and lower efficiency and recovery rates (Chesnot & Schwartzbrod, 2004).

## 4. Conclusion

This study highlights how the selection of concentration, DNA extraction and qPCR quantification methods significantly affect the reported recovery rates of *Cryptosporidium* in wastewater samples. Although great variability exists, most of these methods would still be acceptable for wastewater surveillance efforts. For example, centrifugation demonstrated the greatest recovery among concentration methods and is a low-cost, simple, and easily accessible method for wastewater concentrations. However, its requirement of handling and potential shipping of relatively large volumes of wastewater poses significant challenges for its integration into existing wastewater surveillance efforts and high-throughput workflows.

Meanwhile, filtration through electronegative membranes, a widely adopted approach in wastewater surveillance protocols, produced acceptable recoveries with the addition of a PBST wash. Similar recoveries were also observed for the Nanotrap Microbiome particle protocol, another common wastewater surveillance concentration protocol that is amenable to automated high throughput workflows. To the authors’ knowledge, this study is among the first to evaluate the utility of the Nanotrap Microbiome particles in detecting *Cryptosporidium* in wastewater. Additionally, of the concentration protocols tested, only the IMS purification protocol is not recommended due to unacceptable recovery rates.

Similarly, the choice of DNA extraction kit seemed less critical in affecting DNA yields. A much more important choice was the use of a physical pretreatment prior to DNA extraction. Specifically, bead-beating pretreatment outperformed the freeze-thaw protocol in yielding higher DNA concentrations, likely due to the harshness of the freeze-thaw cycles on DNA integrity.

Thus, as wastewater surveillance expands to other pathogens, including *Cryptosporidium* spp., there are many acceptable concentration, DNA extraction, and qPCR quantification protocols that can be used to fit the unique needs, budgets, and capabilities of each wastewater surveillance effort. However, this study demonstrates that the choices made in the methodology of the wastewater surveillance protocol can have significant effects on the *Cryptosporidium* spp. concentrations reported. Therefore, it is critical to evaluate and report recovery rates to optimize the surveillance protocol and enable more accurate cross-comparisons between studies.

## Data Availability

All data produced in the present study are available upon reasonable request to the authors

## 5. Acknowledgements

We thank the City of Corvallis Wastewater Treatment Facility, Casey Kanalos and James Gallagher for providing and collecting wastewater samples. We thank Ana Child and Gregory Sturbaum of the Portland Water Bureau for their input on methodology selection. We also thank Daniel Goldfarb, Lauren Sanders and Tara Jones-Roe of Ceres Nanosciences, Inc. and Terry Versaw of Streck for their guidance and assistance in using the Ceres Nanotrap Microbiome Particle B workflow. This work was funded by the Centers for Disease Control and Prevention (cooperative agreement no. CK-19-1904).

## Notes

### Competing Interest Statement

The authors have declared no competing interest.

